# Assessing the performance and suitability of wastewater based-surveillance for SARS-CoV-2 RNA in public schools

**DOI:** 10.1101/2025.01.16.25320681

**Authors:** Nicole Acosta, Alex Buchner Beaudet, Paul Westlund, Jia Hu, Navid Sedaghat, Puja Pradhan, Lawrence Man, Jordan Hollman, María A. Bautista, Barbara J. Waddell, Janine McCalder, Matthew Penney, Jianwei Chen, Jon Meddings, Gopal Achari, M. Cathryn Ryan, Dany Breton, Elizabeth A. Wood, Jason L. Cabaj, Rhonda C. Clark, Kevin Frankowski, Casey R.J. Hubert, Michael D. Parkins

## Abstract

Municipal wastewater-based surveillance (WBS) programs for SARS-CoV-2 were valuable tools for epidemiological modelling and informing COVID-19 health policy during the pandemic. We conducted a “near to source” study to assess the capacity and performance of SARS-CoV-2 WBS programs in schools relative to municipal wastewater treatment plants (WWTP). Only 4/17 schools screened had plumbing systems that were amenable to WBS. From December 2020 - March 2021 composite wastewater collected 2X/week from four schools and three municipal WWTP were assessed for SARS-CoV-2 and fecal biomarkers. Schools had lower rates of successful sample collection relative to WWTP (44.7% vs 100%, p<0.001). In a time of low COVID-19 activity, 13/64 of school samples were positive of SARS-CoV-2-N1 vs 100% of WWTP. SARS-CoV-2-N1 detected in school wastewater associated with and preceded clinically identified infections, but did not correlate with rates of overall absenteeism. Notably levels of SARS-CoV-2-N1 and fecal biomarkers were markedly lower in wastewater from schools relative to WWTP. While our SARS-CoV-2 WBS program in schools did generate a leading signal relative to clinical disease, the significantly lower levels of SARS-CoV-2 and fecal biomarkers suggests that toileting habits of children who may avoid defecation at school adversely impact school-based WBS of targets shed in feces.

## Introduction

A silver lining of the COVID-19 pandemic has been the rapid evolution of healthcare innovation to meet the needs of a global pandemic. One technology in particular, wastewater-based surveillance (WBS), has proven especially useful in tracking and understanding the spread of disease ^1^. Wastewater measured SARS-CoV-2 RNA strongly correlates with clinical case burden across a range of scales, acting as an early warning system to inform health policy ^2,3^. Indeed, WBS serves as leading indicator (4-6 days) to clinically confirmed COVID-19 disease ^4^, and strongly correlates with hospitalizations, ICU admissions and deaths across whole communities. This approach is viable because SARS-CoV-2 RNA is shed in the stool of most symptomatic and asymptomatic infected individuals very early in the course of infection ^5^.

WBS programs for SARS-CoV-2 have evolved to become a *de facto* standard and now exist in almost every major municipality, leveraging samples collected at local wastewater treatment plants (WWTPs) to understand community disease patterns^6^. More granular monitoring programs, focused on individual neighborhoods have similarly demonstrated strong correlations with clinically diagnosed cases – with the added potential of understanding community factors related to COVID-19 ^7–10^. More granular yet, “near to source” WBS focuses on specific buildings and facilities to establish the limits of this technology ^11,12^. Areas that were initially explored include those such as hospitals ^13,14^ and long-term care facilities ^15,16^, where the consequences of outbreaks are profound for both the higher risk resident populations and for the ability to disrupt the care these critical services provide to the broader community. One area that has been explored only to a limited degree is the potential for close to source monitoring in public schools.

COVID-19 transmission in pediatric and adolescent populations represents a contentious topic in epidemiological research ^17–20^. Early works concluded that children were responsible for less than 5% of total COVID-19 cases in general populations ^21^. However, the tendency for children and adolescents to exhibit asymptomatic or pauci-symptomatic disease meant that cases were likely to be initially under reported ^17,22,23^. Later studies have since demonstrated that children are a main source of viral transmission within family clusters ^24^. Schools increase transmission potential between unrelated families due to the concentration of students from different households.

In this study, we initiated a ‘near to source’ pilot program, exploring the ability and viability of SARS-CoV-2 WBS to detect COVID-19 in public schools and assessed this relative to overall community disease activity as measured at municipal WWTP. To achieve this aim, we first developed a framework for screening potential schools to identify suitable candidates. Schools that met the required framework criteria then enrolled in a four-month observational study whereby SARS-CoV-2 RNA measured in wastewater was compared with clinically identified cases within the school, and across the broader community. As SARS-CoV-2 is merely the tip of the iceberg for potential WBS measured analytes of public health importance (i.e. other communicable infections, drugs and toxins) understanding how it can perform (and underperform), in schools is essential.

## Results

### Selection of Participating Schools and Wastewater Collection Performance

At the end of 2020, we undertook a study to assess the feasibility of schools to participate in a longitudinal WBS program for SARS-CoV-2. Of the 17 schools that were screened, ultimately only four schools met all of the criteria for a site based near-source WBS pilot program (Supplementary Fig. 1). Detailed characteristics of the four selected schools are included in Supplementary Table 1. Between December 2020 and March of 2021, four schools were sequentially onboarded with the goal of achieving at least three months of sampling from each school. The timing of the study coincided with the end of ‘wave-2’ in Alberta, and was characterized by the wild type SARS-CoV-2 variant ^25^. Of the total number of samples that were sought for collection (i.e. during regular school days), 76.5% (64/79) were successfully collected from four schools (five sites) during the study. As schools were sequentially brought online, the number of samples available from each site differed. Unsuccessful collection of a composite wastewater sample was attributed to a range of issues including ragging (3/15 failures), low sanitary flow (9/15 failures), and ambient temperature excursions (3/15 failures). The occurrence of these events did not occur disproportionally at any specific site (p=0.140, Fisher’s exact test) (Supplementary Table 1). During this time 66/66 (100%) twice weekly samples were collected in parallel from the three municipal WWTPs. Successful collection occurred at a higher rate in municipal WWTPs than schools (100% vs 44.7%), p<0.001. WWTPs samples were not subject to ragging, low sanitary flow, or ambient temperature excursions.

### Identification of SARS-CoV-2 RNA signal in school wastewater and its relation to overall community activity

Only thirteen wastewater samples from schools (of 64; 20.3%) (Table 1) were positive for the SARS-CoV-2 N1 target. Six of the 64 school wastewater samples (9.4%) were also identified as positive for the SARS-CoV-2-N2 target, and no sample positive for N2 was not also positive for N1. School samples that were positive for N2 had higher amounts of SARS-CoV-2 N1 RNA as indicated by a lower cycle threshold (Cq) value (37.8 vs 40, p=0.0003). The median Cq value for positive samples was 37.8 (IQR 36.6-39) for the N1 target and 40.6 (IQR 40.1-42) for the N2 target. Conversely, the lowest Cq value observed was 34.7 and 39.1 for N1 and N2 respectively. A Spearman correlation test between Cq of the N1 and N2 RT-qPCR assays within the same wastewater samples yielded a strong positive correlation (r=0.632, p<0.0001).

**Table 1.** SARS-CoV-2 positive wastewater samples identified from schools ^a^.

| Site Name | Number of students with Confirmed COVID-19 Diagnosis | Number of WW Samples Positive for SARS-CoV-2 RNA | Date of Positive SARS-CoV-2 RNA Reading (DD-MM-YYYY) | Mean N1 Detected Copies/mL | Mean N2 Detected Copies/mL |
| --- | --- | --- | --- | --- | --- |
| **School #1** | 3 | 3 | 01/02/2021 | 4.84 | 0 |
|  |  |  | 10/02/2021 | 9.24 | 3.46 |
|  |  |  | 01/03/2021 | 0.72 | 0 |
| **School #2A** | 11 | 7 | 13/01/2021 | 1.09 | 1.08 |
|  |  |  | 18/01/2021 | 0.401 | 0.945 |
|  |  |  | 20/01/2021 | 0.497 | 0 |
|  |  |  | 22/02/2021 | 7.14 | 9.29 |
|  |  |  | 24/02/2021 | 5.38 | 1.07 |
|  |  |  | 22/03/2021 | 8.94 | 1.91 |
|  |  |  | 24/03/2021 | 25.9 | 0 |
| **School #3** | 3 | 1 | 24/02/2021 | 3.58 | 0 |
| **School #4** | 1 | 2 | 08/03/2021 | 0.478 | 0 |
|  |  |  | 10/03/2021 | 0.601 | 0 |

<sup>a</sup> Calgary, Alberta, Canada wastewater samples collected from four different schools over study period. Each site was monitored for a different timespan within the study dates. Wastewater samples identified as positive for SARS-CoV-2 RNA using N1 and N2 loci RT-qPCR assays are shown (see methods for definition). School #2B not included due to lack of any positive wastewater samples.

To understand SARS-CoV-2 burden in schools relative to the larger community from which they derived, we compared school wastewater data to that across all of the city of Calgary’s via samples collected at WWTPs. During the study period, 66 twice-weekly samples were collected from Calgary’s three WWTP (23/per). All WWTPs samples were positive for SARS-CoV-2 N1 target and all but one for N2. Compared to schools, municipal wastewater was much more likely to be positive for SARS-CoV-2 N1 (66/66 (100%) vs 13/64 (20%), p<0.0001). Furthermore, school SARS-CoV-2 RNA levels were significantly lower than WWTP at a time of low community activity regardless of the target that was evaluated: N1; median Schools 0 (IQR: 0-0) copies/mL vs median WWTP 126 (IQR: 67.8-202) copies/mL and N2; median Schools 0 (IQR: 0-0) copies/mL vs median WWTP 66.8 (IQR: 29.2-131) copies/mL for (Fig. 1A). Similar trends were observed when SARS-CoV-2 concentration was normalized by the PMMoV signal in the samples to potentially control for fecal burden: N1; median Schools 0 (IQR: 0-8.5 x 10^-5^) copies/copies vs median WWTP 1.2 x 10^-2^ (IQR: 6.3 x 10^-3^ – 2.4 x 10^-2^) copies/copies, p<0.0001 and N2; median Schools 0 (IQR: 0-0) copies/copies vs median WWTP 4.8 x 10^-3^ (IQR: 2.1 x 10^-3^ – 2 x 10^-2^) copies/copies, p<0.0001 (Fig. 1B). Even when we excluded those school samples that were negative for SARS-CoV-2 RNA for either the N1 or N2 target, both raw school data and normalized data were markedly lower than WWTP (data not shown).

**Figure 1.**
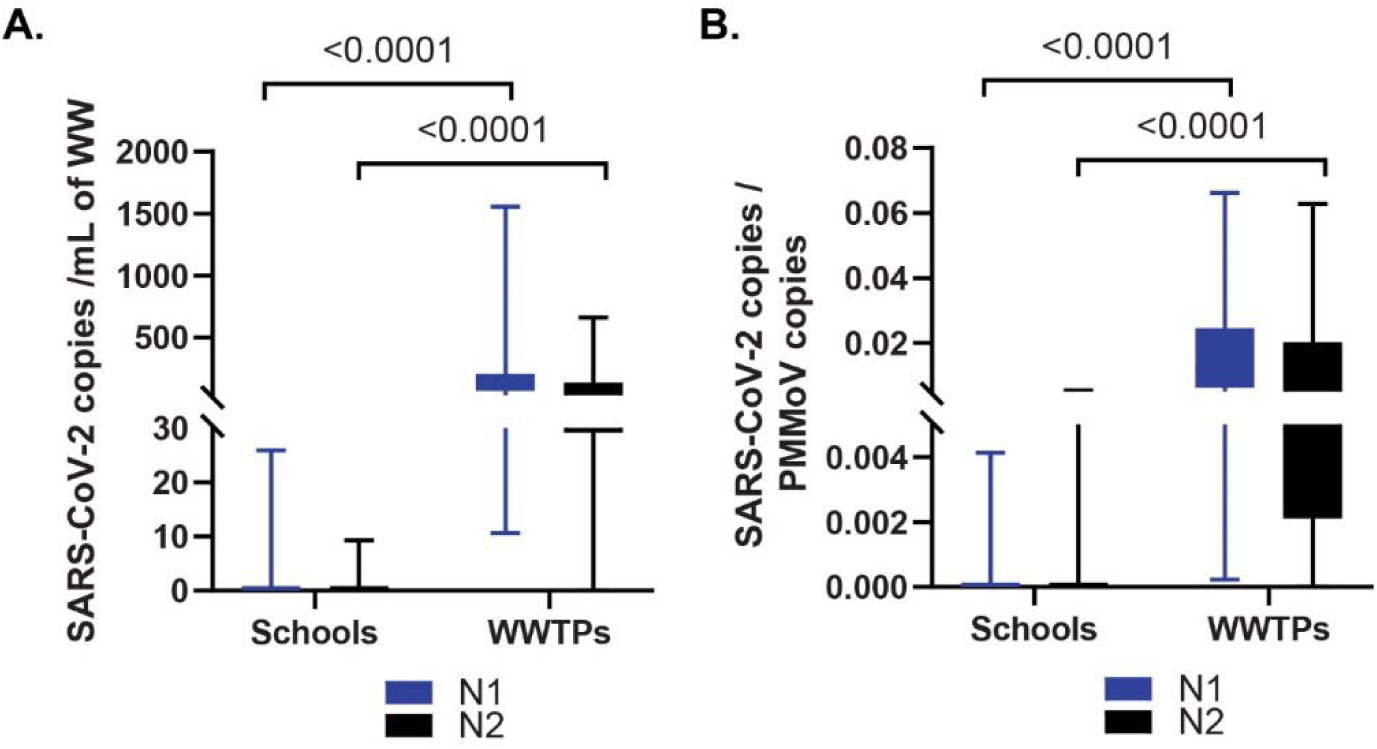
SARS-CoV-2 RNA in wastewater from schools is lower than that from the surrounding community. Comparison of the SARS-CoV-2 RNA signal found in wastewater from schools and WWTP that were positive for SARS-CoV-2 RNA between January to March 2021. Quantification of SARS-CoV-2 RNA in samples was determined by the N1 (blue) and N2 (black) assays. (A) SARS-CoV-2 RNA signal (copies/mL). B) SARS-CoV-2 genomic copies normalized to genomic copies of the fecal biomarker PMMoV. Differences were determined using the Mann Whitney test. WW: wastewater, WWTP: wastewater treatment plants.

### Clinical Case Information

Demographics of the included schools are presented in Table 2. Clinical case information on COVID-19-confirmed, related- and unrelated-infections and overall absenteeism were collected from January 2021. The size of the student and staff population of each school varied throughout the monitoring period (Table 2) as exposures, cases (prompting exclusions) and other illnesses affected their populations (Supplementary Fig. 2). Between January 2021 and March 2021, only eighteen clinically confirmed COVID-19 cases were documented amongst students and staff at the four participating schools in Calgary. The weekly absenteeism rates (due to confirmed COVID-19, COVID-19 related or unrelated illnesses) ranged from 0 to 0.44%, 0.04 to 27.3% and 0.19 to 16.7%, respectively (Supplementary Table 2). Absenteeism rates due to COVID-19 confirmed disease did not significantly change through the three-month study period (p=0.290, Kruskal-Wallis test). Confirmed cases represented a very small proportion of student absences [median percent of COVID-19 confirmed 0.09 (0-0.183) vs any other non-COVID-19 reason 9.32(7.94-12.3), p <0.0001], Wilcoxon matched-pairs signed rank test) on account of the rigorous exclusion policy intended to reduce secondary spread ^26–28^.

**Table 2.** Demographics and characteristics of schools monitored in the city of Calgary.

| **School** | **#1** | **#2** | **#3** | **#4** |
| --- | --- | --- | --- | --- |
| **Category (Grades)** | Elementary<br>and Middle<br>School (K-9) | High School<br>(10-12) | High School<br>(10-12) | High School<br>(10-12) |
| **Daily Student**<br>**Population (range)** | 552-779 | 1158-1335 | 1285-1364 | 1354-1558 |
| **Daily Staff population**<br>**(range)** | 69-78 | 90-103 | 104-110 | 100-108 |
| **Average Attendance**<br>**Rate (%)** | 87.0 | 92.0 | 86.5 | 92.3 |
| **Wastewater Treatment**<br>**plant catchment** | WWTP 1 | WWTP 1 | WWTP 1 | WWTP 2 and<br>3 |

WWTP: wastewater treatment plant

### SARS-CoV-2 RNA in school wastewater and its relation to clinical case burden

Comparative analysis of wastewater measured SARS-CoV-2 RNA and clinically confirmed cases in schools started in January 2021 using complementary approaches. We looked to determine if there was a correlation between a positive SARS-CoV-2 N1 RNA signal (Cq<40) measured in a specific school and the occurrence of clinically confirmed cases in the same school over the ensuing days. We were able to confirm a statistical correlation using the Fisher’s exact test between SARS-CoV-2 RNA detected in school wastewater and clinical cases that were confirmed in the subsequent week (Table 3). Whereas this was statistically significant when wastewater samples were collected at intervals longer than 1-3 days preceding clinical cases, there was only a trend at shorter intervals preceding clinical case occurrence.

**Table 3.** SARS-CoV-2 RNA signal detected in school wastewater and the occurrence of confirmed COVID-19 cases in the following week.

|  | **Confirmed Cases**<br>**and a positive**<br>**WBS-signal** | **Confirmed Cases**<br>**and a negative**<br>**WBS-signal** | **Risk Ratio** | **P-value** |
| --- | --- | --- | --- | --- |
| **1 – 2 days** | 6/13 (46%) | 15/51 (29%) | 1.57 (0.76-3.24) | 0.205 |
| **1 – 3 days** | 9/13 (69%) | 15/51 (29%) | 2.35 (1.34-4.12) | **0.011** |
| **1 – 4 days** | 9/13 (69%) | 16/51 (31%) | 2.21 (1.28-3.80) | **0.015** |
| **1 – 5 days** | 10/13 (77%) | 18/51 (36%) | 2.18 (1.35-3.51) | **0.008** |

In 9/13 incidents where a school wastewater sample was identified as positive for SARS-CoV-2 RNA N1 signal (Fig. 2), another positive sample followed for the next closest sampling date 9/13 (69.2%) vs 4/53 (7.55%) (P< 0.0001, Fisher’s exact test). In the other four positive samples, the sampling dates immediately adjacent to the positive sample date did not have sufficient sample volume collection for analysis, therefore we cannot comment on the trend. Given the exclusion of students once COVID-19 was confirmed, this may represent asymptomatic secondary cases identified through WBS.

**Figure 2.**
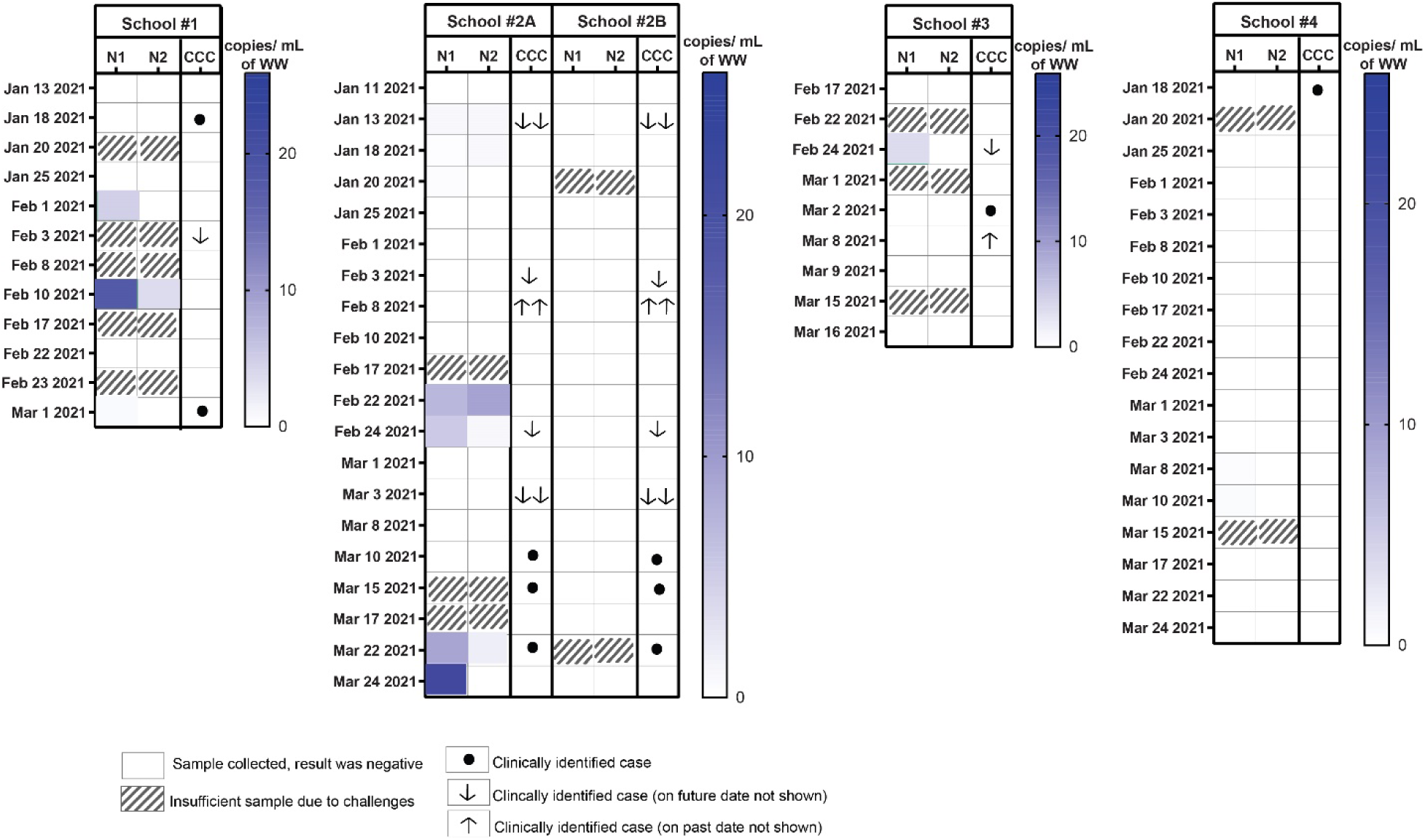
Raw SARS-CoV-2 RNA detection in school wastewater. SARS-CoV-2 N1- and N2-RNA detected are plotted using a heat map. Clinically confirmed cases (CCC) reported to school administrators are graphically shown for each school. An arrow in the CCC columns refers to a clinically identified case on a date other than that of sample collection. The number of arrows indicates the number of days removed from the date indicated (i.e., 2 downward facing arrows on January 13^th^ indicates a clinical case on January 15^th^).

We then sought to understand if overall attendance (i.e., students and teachers) amongst the four schools correlated with wastewater detected SARS-CoV-2 RNA in school wastewater. When we sought to determine if correlations between overall absenteeism (assessed at >5% and >10%) at any individual school with the likelihood of having a positive SARS-CoV-2 N1 signal (Cq<40) in that school using the Fischer’s exact test showed interdependence between absenteeism and positive SARS-CoV-2 signal. No evidence of a statistical association was found at any of the four schools measured through five sites; School #1, School #2A, School #2B, School #3, and School #4 when school absenteeism was >5% (p>0.999, p=0.088, p>0.999, p>0.999, and 0.625 respectively) or >10% (p=0.429, p=0.516, p>0.999, p>0.999, and p=0.375 respectively).

Finally, we sought to better understand school attendance at each site in the context of overall community viral load as determined from a city-wide N1 SARS-CoV-2 signal across each of Calgary’s three wastewater treatment plants (Fig. 3A). Using Spearman correlation test we did not observe a correlation between overall attendance (within 2 days, owing to the frequency of sample collection) for any individual school and the SARS-CoV-2 N1 city-wide aggregate wastewater value for the City of Calgary (Fig. 3B) (Spearman’s r = 0.136, CI: -0.419 -0.616, p= 0.629).

**Figure 3.**
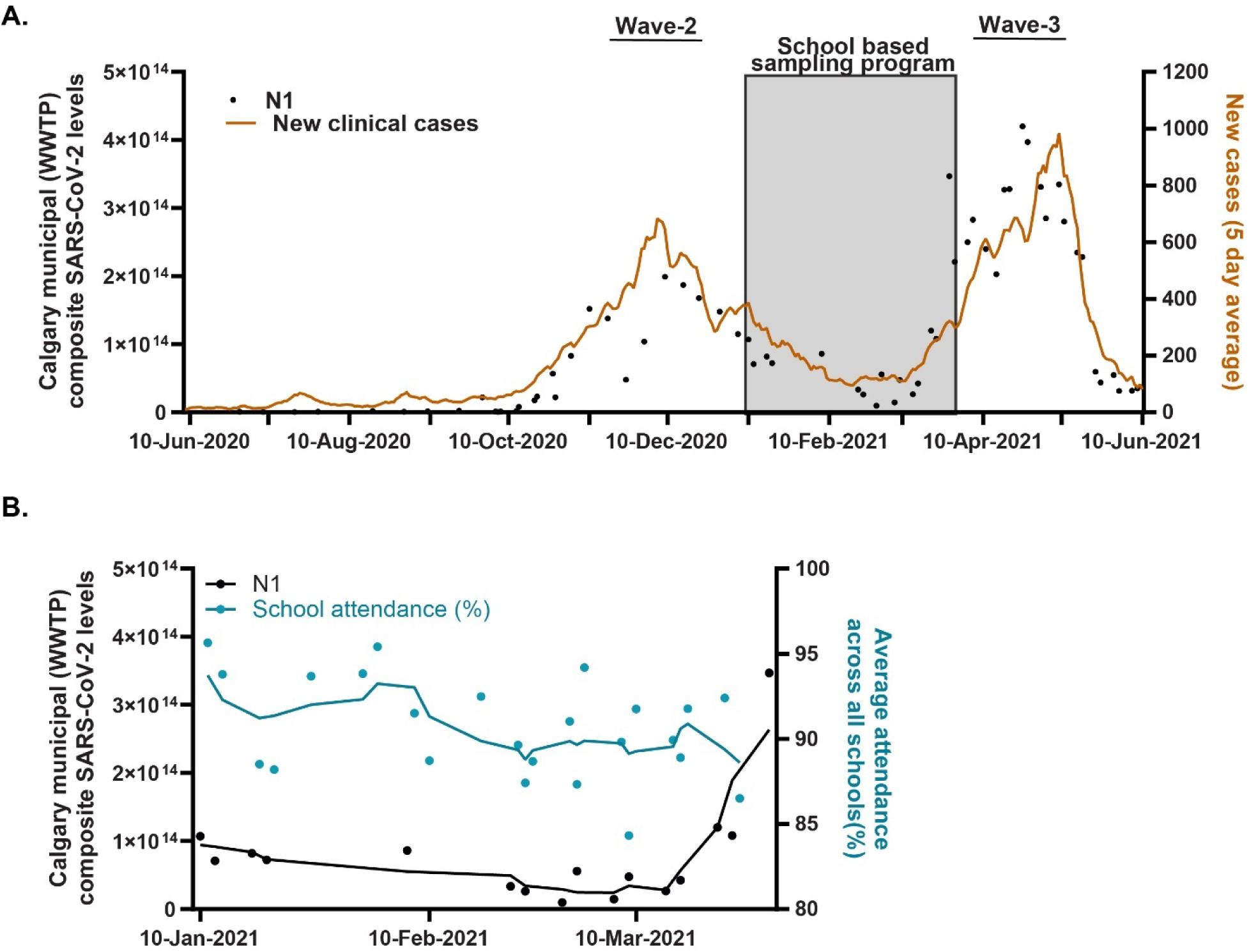
Total mass flux of SARS-CoV-2 RNA across the city of Calgary relative to attendance patterns in the four individual schools monitored by WBS. (A) Daily dynamics of the mass flux of SARS-CoV-2 RNA N1 across the entire city of Calgary from June 2020 to June 2021 using the N1 (black dots) assay. Clinically confirmed COVID-19 new cases (5-day average) are represented as orange lines. The sampling period for the four schools is highlighted in grey – occurring between waves 2 and 3. (B) Average composite attendance (staff and students) across all four schools compared against community burden of SARS-CoV-2 as measured in Calgary’s wastewater. This figure serves as an assessment of the correlation between the total SARS-CoV-2 wastewater-N1 (across all of Calgary) with the overall attendance percentage (within 2 days, owing the frequency of sample collection) for all 4 schools participating in the program using the Spearman correlation test. Lines represent the best fit plotted with second order smoothing for the N1 SARS-CoV-2 concentration (copies per day) (Black) and average school attendance across the four monitored sites (%) (Blue) from January to March 2021. WBS: wastewater-based surveillance, WWTP: wastewater treatment plants.

### Unique Features of School-based WBS

Sources more proximal in the sewershed have greater potential for molecular inhibition from chemical and other molecular factors. To assess if differences might explain our data, we compared recovery of spiked-BCoV into wastewater from the schools and WWTPs samples. There was a small but statistically significant difference in the spiked external positive control BCoV median copy numbers between schools and WWTPs (694,914 [IQR: 55,703-1,675,777] copies/mL vs 1,217,451 [IQR: 774,211-1,934,018], p=0.005, Mann Whitney test) (Supplementary Fig. 3A). However, school 2B was a significant outlier and had significantly lower detection (∼10,000-fold) of BCoV (Supplementary Table 3) suggesting the potential for substantial signal interference (Supplementary Fig. 3B). When we removed site 2B as a significant outlier, school median BCoV did not differ from the WWTPs (Supplementary Fig. 3C).

To assess for differences in fecal contributions at each school, and therefore the potential to detect fecal shed WBS targets relative to the community at large we took multiple complementary approaches. The human fecal biomarker, PMMoV, was much lower in school samples than in WWTP (School median 2,178 [IQR: 28.1 – 8,213] copies/mL vs WWTP median 13,029 [IQR: 5,489 – 19,556], p<0.0001, Mann Whitney test) (Supplementary Fig. 4A-B). As school site 2B remained a significant outlier (Supplementary Table 3), we analyzed this separately as well with the same trends observed (School median 4054 [IQR: 1,853 – 11,092] copies/mL vs WWTP median 13,029 [IQR: 5,489 – 19,556], p=0.0003, Mann Whitney test) (Supplementary Fig. 4C). When 2B site was excluded, the median school PMMoV burden remained 3.2-fold lower than the average WWTP, suggesting a lower fecal content. Chemical analysis demonstrated that total suspended solids (TSS) and total volatile suspended solids (TVSS) in wastewater from schools was significantly lower than WWTP (Supplementary Fig. 5).

## Discussion

To understand the potential for near-source WBS in schools, we analyzed the spatial and temporal differences among SARS-CoV-2 among four large public schools in the city of Calgary. The data collected in this study suggests that, despite the sporadic patterns of wastewater flow in individual buildings of visitation such as schools, the presence of SARS-CoV-2 RNA can be identified. Recently groups have confirmed that the incidence of SARS-CoV-2 in wastewater of schools, corresponds with the overall community disease burden ^12,29–31^. Our group took this one step further and compared clinical disease occurring in student/staff population – even at a period of low COVID-19 community transmission – with WBS, identifying that indeed this signal correlated with WBS. By augmenting site specific-WBS with an overall community SARS-CoV-2 WBS through municipal WWTP enables a comprehensive, holistic perspective. This complements recent publications from other school-based WBS programs from countries around the world including UK, USA, Czech Republic, Canada, and Thailand^12,29–36^. Some of those studies found that the amount of SARS-CoV-2 RNA signal in school wastewater correlated with the number of incident COVID-19 cases ^30,31^, and in some instances this provided a leading signal ^12,33,36^. In contrast, other school WBS studies reported only moderate correlations between SARS-CoV-2 RNA in wastewater and clinical cases of COVID-19, specifically during periods of low COVID-19 prevalence^34^. Here we identified during a period of low community activity that WBS, performed twice weekly, provided a 1-3 to 1-5 lead time relative to clinical case diagnoses. This is similar to Kappus-Kron et al., who identified a one day lead time relative to clinical diagnoses in a weekly sampling program at a school of ∼600 students in New York State ^36^.

Interestingly, WBS in other near source settings such as colleges, university campuses and residence halls have generally shown a stronger relationship with clinical diseases in those sites ^37–42^. One likely explanation for this discordance likely relates to human factors such as toileting ^7,33,43^. As SARS-CoV-2 is almost exclusively shed in the stool ^44^, successful monitoring is dependent on toileting behavior of the population using the facilities. This is a critical barrier for WBS programs in schools. An average of 63% of students between the ages of 6 and 16 years refuse to defecate at schools^45^ and this avoidance behavior is most pronounced in high school students. This is supported by significantly lower evidence of fecal biomarkers including PMMoV, TSS and TVSS observed in schools (and in particular high schools) relative to community signals overall. Unfortunately, this limits the utility of school WBS programs for fecal shed targets – such as SARS-CoV-2– and represents a novel observation in this study. This would therefore likely impact pathogens such as Influenza, Respiratory Syncytial Virus (RSV), and viral causes of gastroenteritis but not measles and other viruses shed in high amounts in urine, which have all been previously identified in sewage ^46–49^.

Importantly, school-based WBS applications come with significant technical challenges. Only 4/17 (24%) of schools screened for potential participation had infrastructure that enabled a comprehensive program monitoring the entire school from ≤2 sites while excluding wastewater from other community inputs. While multiple complementary devices could in many instances overcome this short coming, they would result in significant increase in the work and costs associated with WBS ^33^. Once installed, school-WBS devices did not successfully procure samples at the same rates as WWTP and were more likely subject to complications requiring specific maintenance (i.e. ragging and blocking). These challenges have been observed in other near to source programs and highlight how fortunate municipal based monitoring programs are to have experts on site for autosampler maintenance and sample management.

Several limitations of note are to be considered in regard to this work. This study was conducted during the trough of Wave-2 - at the end of the 2020-2021 winter season and early spring of 2021 ^50^, during a time of very low overall community COVID-19 activity^7^. The low incidence were in response to the strict public health measures (included province-wide mask mandate, encouragement of work-from-home, and prohibition of social gatherings ^51^) that were implemented in order to resume in-person learning in schools after the winter holiday break. During higher incidence times, the performance may have been different. While composite samples were collected differently for schools vs WWTP (8 vs 24-hours), the limitation of school WBS to hours of attendance would have increased its potential to capture defecation events, which it did not.

## CONCLUSION

SARS-CoV-2 WBS performed in schools is capable of identifying incident cases of COVID-19 before they occur. However, significant limitations exist. Most schools do not have a plumbing network that enables single/dual source surveillance, and sample collection is challenging relative to conventional WWTP programs. Furthermore, as students are less likely to defecate at school, the capacity for this technology to effectively be used for SARS-CoV-2 and other fecal shed WBS targets is reduced.

## Methods

### Public school and community wastewater sampling

In partnership with the Alberta Health Services (AHS) Medical Office of Health, and a Calgary-based school board, we first sought to identify candidate schools potentially amenable to WBS. Provisional inclusion criteria included a population of ≥500 students and a willing administration. The process of identifying appropriate sampling sites included a series of sequential screenings (Supplementary Fig. 1). Sites were excluded only after failing screening. The first step of screening involved assessing each building’s sewer network mechanical drawings - necessary to determine if plumbing access ports allowed for wastewater collection from all toilets, sinks and other source locations in the building from ≤ two sites, while simultaneously avoiding external sources from the surrounding neighborhoods. The second screening step involved a physical evaluation of potential plumbing access ports. Acceptable sewer access ports could not pose safety hazards or a hindrance to student/faculty activity during installation or ongoing sample collections/maintenance. The final screening was an extensive in-person physical review of internal plumbing of each facility. This involved checking the relevant infrastructure for any obstructions or unavoidable structural hindrances and identifying where wastewater could be comprehensively and safely be collected. If within building collection was not possible, we reviewed municipal access points to see if they could effectively substitute. C.E.C autosamplers were deployed at the sewer access port(s) of each school deemed most appropriate, which are detailed in Supplementary Table 1. Autosamplers were programmed to operated continuously. Collection from school #3 was impeded by a physical obstruction identified after passing all steps, however, the outdoor municipal sewer access port which comprehensively and exclusively served this building was available. A wastewater sampling routine was developed in which wastewater was collected only during times in which students and faculty were present at schools. Details of individual sampler programming is available in the Supplementary Table 1. This study was conducted after receiving approval from the University of Calgary’s Conjoint Health Research Ethics Board (REB20-1544).

To compare the burden of SARS-CoV-2 in school populations relative to the community, raw wastewater from each of Calgary’s three wastewater treatment plants (WWTP) was collected up to three times per week by City of Calgary Water Services staff as previously described^7^. Briefly, ISCO 5800 and ISCO 6712 portable autosamplers were programmed to collect and store 24-hour composite community samples, in which WWTP-1 samples were flow-weighted and samples from WWTP-2 and WWTP-3 were time-weighted to create a single city-wide metric (see below for details)^7^.

School and WWTPs samples were transported to the University of Calgary Advancing Canadian Water Assets (ACWA) laboratory on ice for sample concentration and nucleic acid extraction. Samples that were not successfully collected were categorized based on complication type: ragging (abundance of fibrous material blocked sampling inlet tubing), low sanitary flow, and temperature excursions (defined as freezing of sample or malfunction of sampler due to ambient temperature) detailed in Supplementary Table 1.

### Sample concentration and nucleic acid extraction

Wastewater samples were processed in real-time following a previously described methodology^13^. Briefly, each sample was thoroughly agitated to ensure maximum homogeneity, then an aliquoted of 40 mL was spiked with 200 μl of a Bovine coronavirus (BCoV) exogenous control (final concentration of 2500 TCID50/ml) and then subjected to the sample processing and nucleic acid purification steps of the modified 4S (Sewage, Salt, Silica and SARS-CoV-2) Silica Column purification method ^13,52^. An extraction blank (i.e., UltraPure™ DNase/RNase-Free Distilled Water (Invitrogen)) was included in each batch of processed samples. Extracted nucleic acids were transported on dry ice to the Health Sciences Center, to minimize the potential for contamination and for subsequent molecular analysis. Residual wastewater samples were stored at -20°C before chemical analysis.

### Molecular analysis

Gene targets assessed by RT-qPCR included; the SARS-CoV-2 nucleocapsid gene (N1 and N2), the exogenous control BCoV, and the fecal biomarker PMMoV (a fecal burden biomarker), which were all quantified using a QuantStudio-5 Real-Time PCR System (Applied Biosystems) using the same protocols described previously ^13^. Each RT-qPCR was performed in triplicate as per MIQE guidelines (Bustin et al., 2009). Samples were classified as positive for SARS-CoV-2 if either the N1 or N2 gene targets yielded a threshold quantification cycle (Cq) less than 40 ^13,53,54^. City-wide measurement of SARS-CoV-2 burden (copies/day) in wastewater from Calgary’s three WWTP was calculated as the sum of the mass flux from each of the three WWTPs, where it is the product of the SARS-CoV-2 concentration (C, copies/ml) and the daily volumetric flow ^7^. PCR inhibition was assessed using a spike and dilution method as previously described ^7^ using a representative 40 μL purified nucleic acid sample derived from the school wastewater (School #2B) extracted without the addition of the internal control (i.e., BCoV) and a control sample of Ultrapure™ DNase/RNase-Free Distilled water (ThermoFisher). Wastewater samples with a ≥2-Cq delay relative to the controls were considered to have evidence of RT-qPCR inhibition ^55^.

### Chemical Analysis

Calculation of total Solids (TS), total suspended solids (TSS), total volatile suspended solids (TVSS) and total dissolved Solids (TDS) in a wastewater sample was performed following standard methods ^56^. Briefly, to determine TS, homogenized 50 mL aliquots were placed in pre-weighed crucibles and dried at 104 °C for 12 hours to evaporate liquid content. Then, the residue after the drying process was cooled, weighed at room temperature yielding TS. To determine TSS, homogenized 20 mL aliquots were filtered through 1.5 μm pore size Grade 934-AH® RTU glass microfiber filters (Whatman). Then, the residue retained on the filter was dried at 104 °C for 12 hours and weighed to calculate TSS. To determine TVSS, the residue from the TSS step was dried again at 550°C for two hours to drive off volatile solids in the sample. Then, the residue after the second drying process was cooled, weighed at room temperature and TVSS was calculated. Finally, to determine TDS the filtrate from the TSS step was evaporated and dried at 104 °C for 12 hours, and the residue was weighed. Chemical analysis of the WWTPs samples was performed by the staff of City of Calgary following the same standard methods.

### Clinical case data

All clinically confirmed cases of COVID-19 of students attending each school were identified in real-time by a single Provincial health provider, AHS using established protocols and reported to each school by the Office of the Medical Officer of Health of AHS. Daily aggregate case counts including new/incident cases of COVID-19 in students were shared with the study team. Individuals with confirmed COVID-19 by public health order were excluded from attending school for 10 days after their symptoms began. At the time of the study, if a case was identified in any individual class, all other class members were excluded from attending school for the next 14 days. Daily aggregate attendance counts for student/staff were recorded and daily student/staff enrollment were shared with the study team.

### Statistical Analysis

Spearman correlation tests were used to test the correlation between cycle threshold (Cq) values of N1 and N2 region detection with our RT-qPCR assays within the same wastewater samples. Differences in SARS-CoV-2 RNA N1 & N2, BCoV and PMMoV between schools and communities (WWTPs) were determined using the Mann Whitney test. Dunn’s Multiple Comparison Test was used to determine which specific school sites differed from others for WBS targets. Differences in student absenteeism rates due to confirmed COVID-19 cases versus other reason was determined using the Wilcoxon matched-pairs signed rank test. To compare the student absenteeism percentages due to confirmed COVID-19 cases across the weeks for all the schools Kruskal-Wallis test was performed. Cases confirmed within 1 to 5 days before/after wastewater sample collection were compared using Fischer tests to assess if positive school wastewater associated with confirmed cases in schools. To assess if positive cases were associated with school absenteeism, we assessed the correlation between the SARS-CoV-2 wastewater-N1 (Calgary) with the overall attendance percentage using the Spearman correlation test. Additionally, we conducted a series of comparisons between absenteeism with the likelihood of getting a positive SARS-CoV-2 wastewater result at each school using the Fischer test. Statistical analyses were done using GraphPad’s Prism-10 software.

## Acknowledgements

The investigators are grateful to the administration, staff, students and facility operators of each of the schools screened, and especially those four ultimately participating in WBS pioneering. The investigators are grateful to all staff of City of Calgary and especially Water Services, and the participating school board for their participation and support.

## Data availability

All data generated or analyzed during this study are included in this published article (and its Supplementary Information files).

## Abbreviations

BCoV: Bovine coronavirus
PMMoV: Pepper mild mottle virus
SARS-CoV-2: Severe Acute Respiratory Syndrome Coronavirus 2
WWTPs: wastewater-treatment plants
WBS: wastewater-based surveillance
TS: Total solids
TSS: Total suspended solids
TDS: Total dissolved solids

