## Supplementary Material file for "Assessing the performance and suitability of wastewater based-surveillance for SARS-CoV-2 RNA in public schools"

**SUPPLEMENTARY TABLES**

**Supplementary Table 1**. Characteristics of the school samplers and sampling configurations used for wastewater collection.

| **Site Name** | **School #1** | **School #2A** | **School #2B** | **School #3** | **School #4** |
| --- | --- | --- | --- | --- | --- |
| **Monitoring Device** | In-building, comprehensive small C.E.C. autosampler | **1 of 2** complementary in-building small C.E.C. autosamplers | **2 of 2** complementary in-building small C.E.C. autosamplers | Large Municipal autosampler (Teledyne ISCO 6712) | In-building, comprehensive small C.E.C. autosampler |
| **Sampler Location** | Beside common area | Near auditorium | Nearby main washrooms | * External basement doors (outdoors) | Near gymnasium |
| **Sampling Regiment** | Continuous 100mL/min between 0700h-1600h | Continuous 100mL/min between 0700h-1600h | Continuous 100mL/min between 0700h-1600h | *Composite 100mL/15 min between 0700h-1600h | Continuous 100mL/min between 0700h-1600h |
| **Total ideal sample volume** | 2L | 2L | 2L | 10L | 2L |
| **Collection setup** | 4x 500mL bottles - in series | 4 x 500mL bottles - in series | 4 x 500mL bottles - in series | One 10 L bottle | 4 x 500mL bottles - in series |
| **Sampling Days** | Mondays & Wednesdays | Mondays & Wednesdays | Mondays & Wednesdays | *Mondays & Tuesdays | Mondays & Wednesdays |
| **Details of samples collected** | | | | | |
| **Sampling Date Range** | 07/12/2020 – 01/03/2021 | 11/01/2021 – 24/03/2021 | 11/01/2021 – 24/03/2021 | 17/02/2021 – 16/03/2021 | 18/01/2021 – 24/03/2021 |
| **Number of suitable samples collected** | 7 | 17 | 18 | 6 | 16 |
| **Sampling days excluded because of holidays or other planned closures** | 3 (21.4% of total days) | 2 (9.0% of total days) | 2 (9% of total days) | 0 | 2 (10% of total days) |
| **Failure of Sample Collection** | | | | | |
| **Number of unsuccessful sampling events** | 5 (41.6% of sampling days) | 3 (15% of sampling days) | 2 (10% of sampling days) | 3 (33.3% of sampling days) | 2 (11.1% of sampling days) |
| **Ragging** | 0 | 3 (15% of sampling days) | 0 | 0 | 0 |
| **Low Sanitary flow** | 5 (41.6% of sampling days) | 0 | 2 (10% of sampling days) | 0 | 2 (11.1% of sampling days) |
| **Temperature Excursions** | 0 | 0 | 0 | 3 (33.3% of sampling days) | 0 |

** School #3 was monitored via an outdoor municipal sewer access port because no viable sampler installation location existed within the school that did not impede student traffic flow. The Teledyne ISCO 6712 autosampler at School #3 could not collect continuously. Municipal personnel were not available for sample collection on Wednesdays – and as such samples were collected on Tuesdays to fit into their schedules. Equipoise between the two sampler types has previously been established ^1^.*

***Wastewater sampling from school #2 required two separate sampling locations to comprehensively surveil due to the layout of the building’s sewer network.*

*min=minutes*

**Supplementary Table 2.** Student absenteeism rates from participating schools.

| **Week** | **Absent reason** | **School #1** | **School #2** | **School #3** | **School #4** |
| --- | --- | --- | --- | --- | --- |
| Feb 1-5 | COVID-19 confirmed case ^a^ | 0.20% | 0.04% | 0.21% | 0.09% |
|  | Isolation or Illness/Isolation ^b^ | 4.03% | 2.21% | 3.03% | 1.10% |
|  | Other ^c^ | 9.37% | 8.25% | 9.73% | 5.38% |
| Feb 8-12 | COVID-19 confirmed case ^a^ | 0.11% | 0.10% | 0.26% | 0.00% |
|  | Isolation or Illness/Isolation ^b^ | 3.20% | 13.34% | 12.82% | 0.36% |
|  | Other ^c^ | 19.68% | 12.69% | 18.84% | 9.27% |
| Feb 15-19 | COVID-19 confirmed case ^a^ | 0.00% | 0.00% | 0.04% | 0.00% |
|  | Isolation or Illness/Isolation ^b^ | 0.54% | 3.08% | 10.24% | 0.14% |
|  | Other ^c^ | 1.17% | 0.45% | 0.19% | 0.28% |
| Feb 22-26 | COVID-19 confirmed case ^a^ | 0.00% | 0.07% | 0.11% | 0.10% |
|  | Isolation or Illness/Isolation ^b^ | 0.26% | 4.55% | 7.93% | 9.21% |
|  | Other ^c^ | 7.83% | 9.58% | 13.50% | 8.10% |
| Mar 1-5 | COVID-19 confirmed case ^a^ | 0.11% | 0.07% | 0.05% | 0.00% |
|  | Isolation or Illness/Isolation ^b^ | 2.90% | 7.20% | 11.58% | 2.49% |
|  | Other ^c^ | 7.92% | 10.40% | 13.15% | 7.69% |
| Mar 8-12 | COVID-19 confirmed case ^a^ | 0.13% | 0.40% | 0.13% | 0.00% |
|  | Isolation or Illness/Isolation ^b^ | 4.46% | 27.25% | 11.39% | 0.07% |
|  | Other ^c^ | 8.10% | 8.55% | 13.21% | 8.43% |
| Mar 15-19 | COVID-19 confirmed case ^a^ | 0.31% | 0.50% | 0.05% | 0.00% |
|  | Isolation or Illness/Isolation ^b^ | 0.92% | 20.31% | 5.95% | 0.04% |
|  | Other ^c^ | 8.00% | 10.87% | 14.68% | 9.46% |
| Mar 22-26 | COVID-19 confirmed case ^a^ | 0.43% | 0.44% | 0.13% | 0.00% |
|  | Isolation or Illness/Isolation ^b^ | 5.37% | 6.23% | 8.67% | 5.46% |
|  | Other ^c^ | 8.53% | 11.16% | 17.35% | 11.22% |

^a^ Student or staff were absent due to a confirmed diagnosis of COVID-19 reported to the schools from the Medical Officers of Health.

^b^ Isolation refers to when a student was absent due to close contact with a person who has or is suspected to potentially have COVID-19 and/or returning from travel outside of Canada. Illness/Isolation refers to when a student was presented with one or more of the primary symptoms that require self-isolation.

^c^ Others include different reasons: absent without contact from the legal guardian; absent and the office had been informed by a legal guardian; a family holiday or extended absence; illness; medical appointment; or unavoidable cause.

**Supplementary Table 3.** Molecular target identification of internal controls in monitored schools.

| **Target** | **Site Name** | **School #1** | **School #2A** | **School #2B** | **School #3** | **School #4** |
| --- | --- | --- | --- | --- | --- | --- |
| **BCoV (copies/ml)** | **Median (IQR)** | 1,684,316 (1,377,175- 2,956,614) | 1,421,755 (525,848-2,041,764) | 119.0 (13.38-20,575) | 813121 (350,105-1,426,700) | 801839 (491,216-1,669,363) |
| **PMMoV (copies/ml)** | **Median (IQR)** | 3342 (278.9-11,970) | 3178 (1318-8814) | 0 (0 - 0) | 13,999 (2182-31,455) | 4411 (2488- 18,394) |

**Supplementary Table 4.** Mean of chemical parameters measured in wastewater samples from schools.

| **Site Name** | **TS (mg/L)** | **TSS (mg/L)** | **TDS (mg/L)** | **TVSS (mg/L)** |
| --- | --- | --- | --- | --- |
| School #1 | 584±183 | 161±78.1 | 423±127 | 129.4±69.7 |
| School #2A | 876±238 | 364±182 | 512±169 | 269.2±199.3 |
| School #2B | 233±169 | 6.56±15.0 | 226±171 | 4.3±12.7 |
| School #3 | 1406±320 | 392.6±185.9 | 1013±263 | 371.6±168.0 |
| School #4 | 1603±1715 | 376.0±582.7 | 1229±1547 | 339.5±523.3 |

TS: total solids, TSS: total suspended solids, TDS: total dissolved solids and TVSS: total volatile suspended solids.

**SUPPLEMENTARY FIGURES**

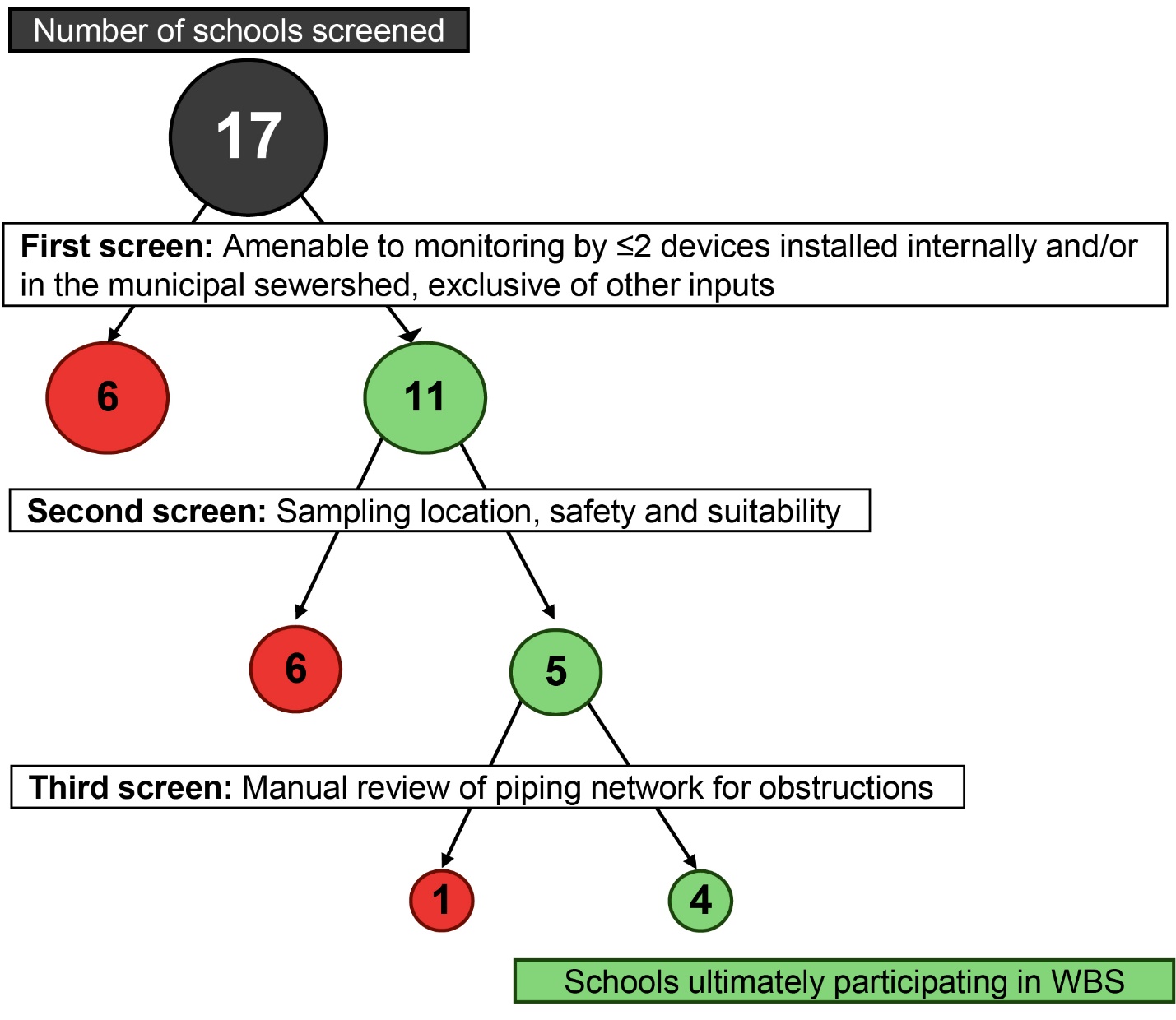

**Supplementary Figure 1**. **School inclusion workflow.** A flowchart of the three-step screening approach used for selecting suitable wastewater sampling locations for school-based WBS. Sites evaluated were primarily within buildings of interest. However, external sewer access ports were evaluated with assistance from a municipal wastewater management team when appropriate outdoor locations were identified. This process ultimately resulted in four (of 17 screened) candidate schools participating in WBS.

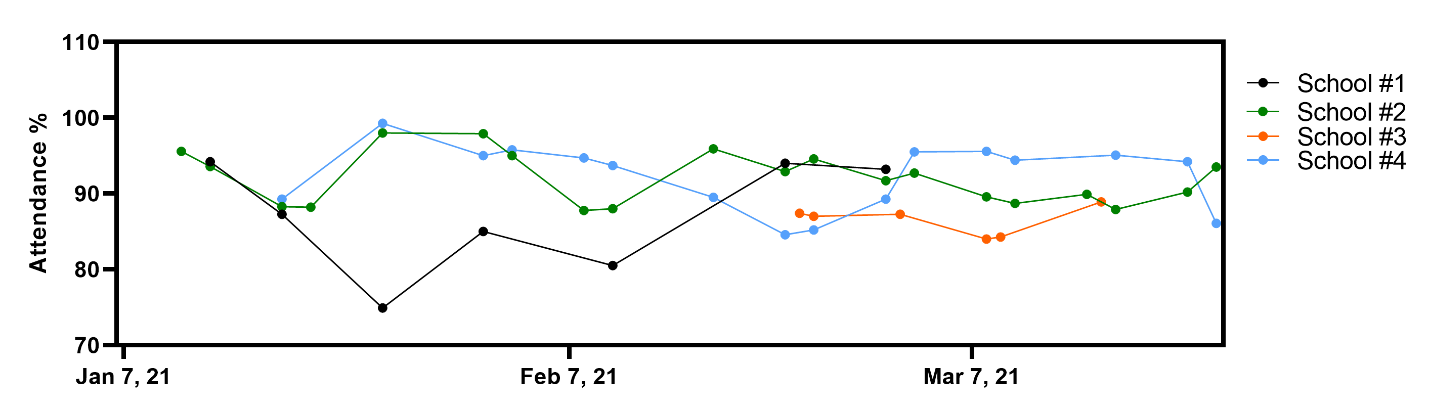

**Supplementary Figure 2.** Aggregate attendance rates of students and teachers for each school under study over the course of the study.

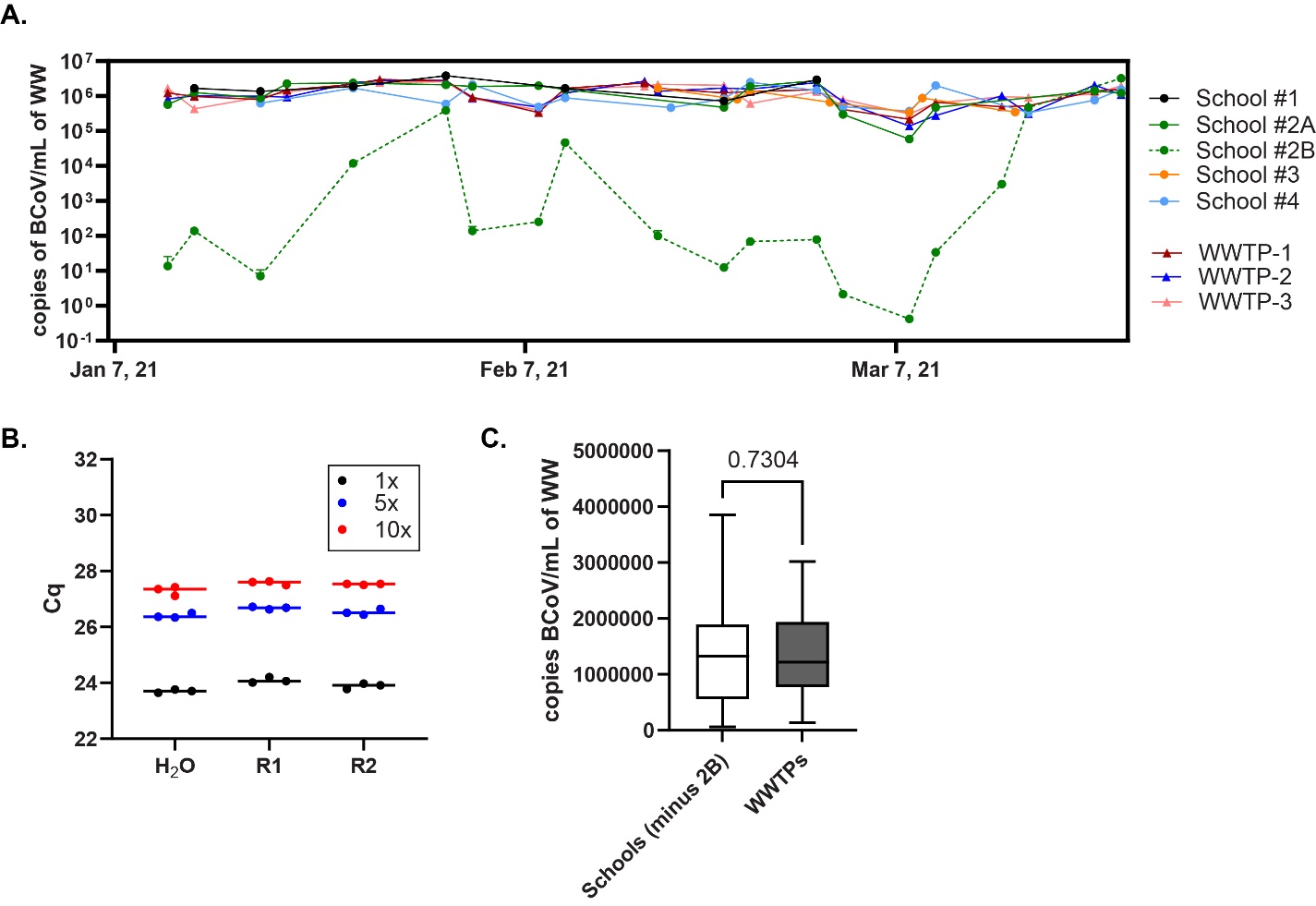

**Supplementary Figure 3. Concentration of Bovine Coronavirus (BCoV) recovered from spiked samples.** (A) BCoV RNA concentration (copies/ml) time course for all schools and wastewater treatment plants (WWTPs) analyzed. Plots show little variability across all sites except for school #2B where considerable variation in recovered BCoV was evident over time. Associated Kruskal-Wallis tests (Dunn’s multiple comparison test) showed a significance difference in mean BCoV concentration between School #2B and all other sites (maximum p=0.0007) besides School #3 (p=0.086). (B) RT-qPCR inhibition determination assay in wastewater samples from School 2B; each of the three sample results show the average of three technical replicates with SD. No RT-qPCR inhibition was observed in the representative School #2B samples. (C) Comparison of genomic copies of BCoV per ml between schools (excluding samples from School 2B site) and WWTP wastewater samples. Differences in BCoV wastewater signal between schools and WWTP were determined using the Mann Whitney test. WW: wastewater, WWTP: wastewater treatment plants, Cq: cycle of quantification.

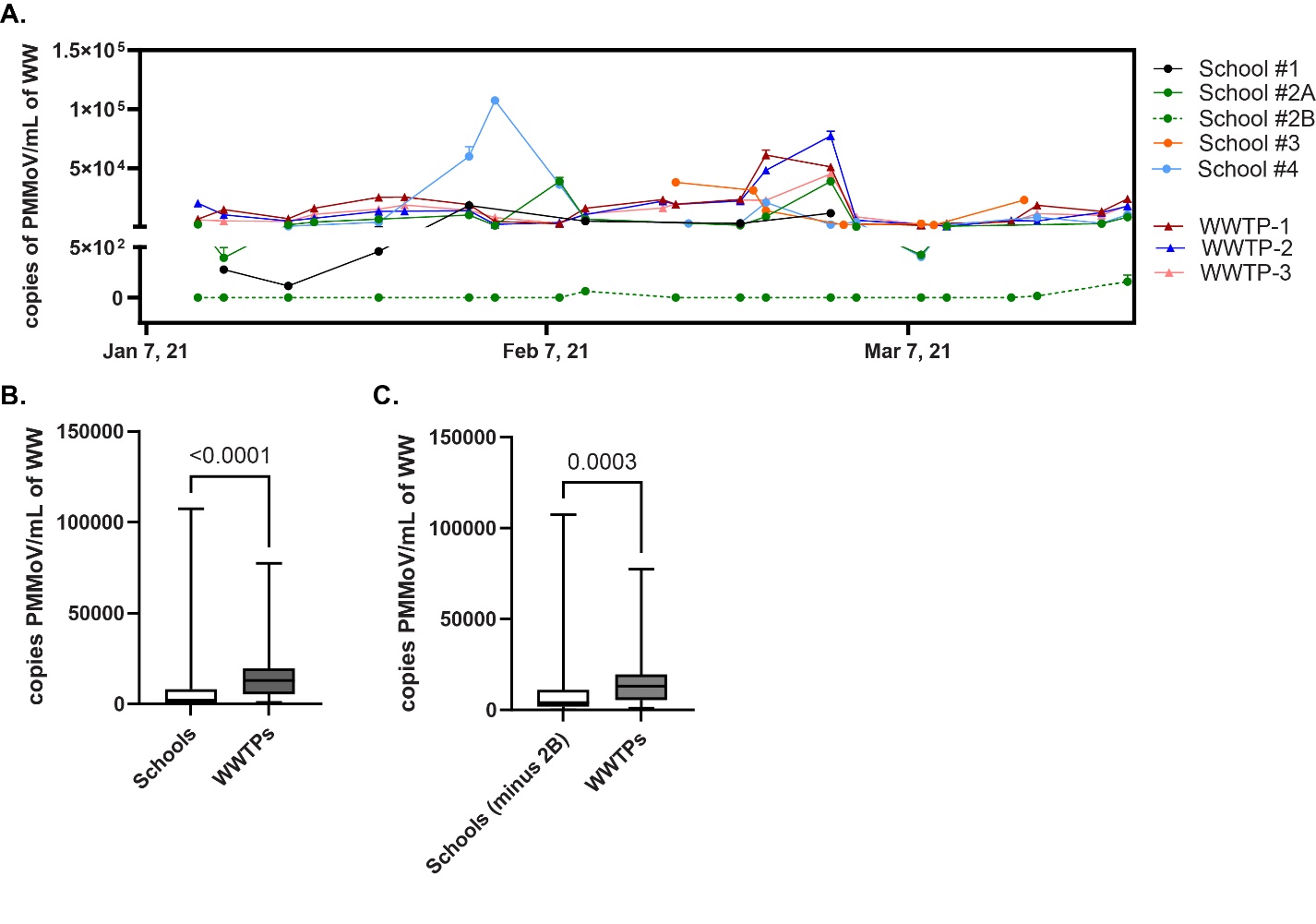

**Supplementary Figure 4.** **Wastewater burden of Fecal Biomarkers is lower in schools.** (A) PMMoV RNA concentration (copies/ml) time course for all 4 schools and wastewater treatment plants (WWTPs) analyzed. Time course plot demonstrates on most occasions the concentration of PMMoV in school wastewater was very low, with only rare large spikes in PMMoV across sites. Associated Kruskal-Wallis tests (Dunn’s multiple comparison test) showed a significance difference in mean PMMoV concentration between School #2B and all other sites (maximum p=0.0094). (B) Comparison of genomic copies of PMMoV per ml of wastewater processed in schools and WWTP wastewater samples. (C) Same data set as panel B excluding samples from School 2B site. Differences in PMMoV wastewater signal between schools and WWTP were determined using the Mann Whitney test. WW: wastewater, WWTP: wastewater treatment plants.

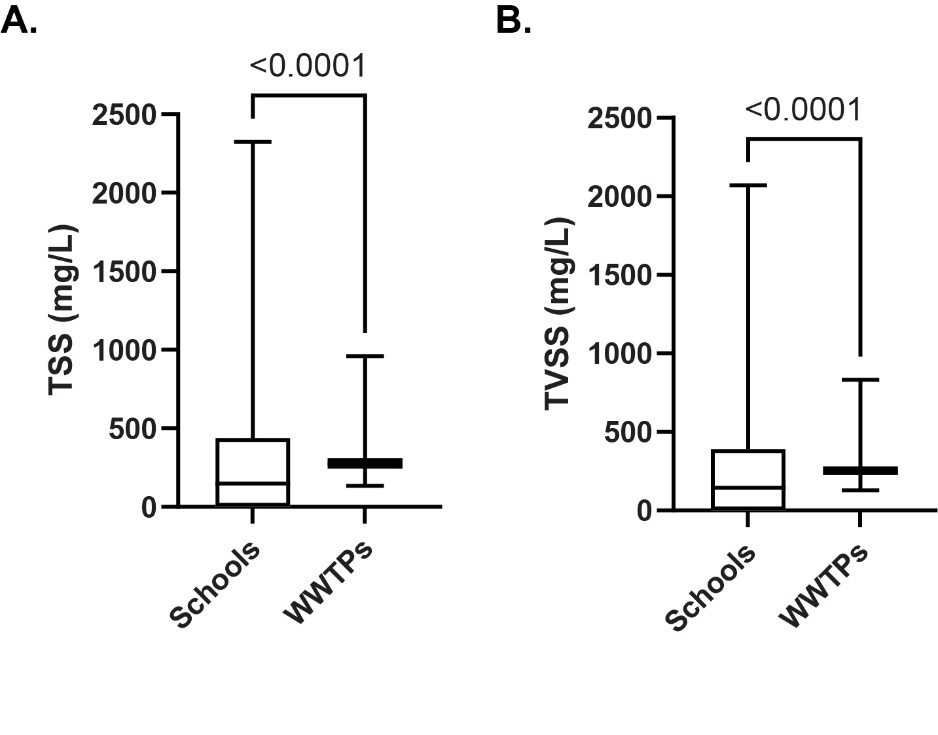

**Supplementary Figure 5. Chemical parameters in school and community wastewater samples.** Differences in chemical parameters between schools and communities (WWTPs) were determined using the Mann Whitney test. Quantification of total suspended solids (TSS) (A) and total volatile suspended solids (TVSS) (D) in school’s wastewater samples vs WWTPs. WW: wastewater, WWTPs: wastewater treatment plants.
